# Gym-Based Modified Powerlifting Exercises for People with Early Onset Parkinson’s Disease: Study Protocol

**DOI:** 10.1101/2024.05.06.24306601

**Authors:** Dale M Harris, Claire Thwaites, Michelle L Callisaya, Richard Blazé, Meg E Morris

**Affiliations:** Institute for Health and Sport (IHeS), Victoria University, Melbourne, VIC 3011, Australia; Peninsula Clinical School, Central Clinical School, Monash University, 2 Hastings Road, Frankston, VIC, Australia; Menzies Institute for Medical Research, University of Tasmania, 17 Liverpool Street, Hobart, TAS, Australia; Epworth Neurosciences Clinical Institute, Epworth, Melbourne, VIC 3121, Australia; Academic and Research Collaborative in Health (ARCH), La Trobe University, Bundoora, VIC 3086, Australia; Healthscope Victorian Rehabilitation Centre, Glen Waverley, VIC 3150, Australia; Care Economy Research Institute, La Trobe University, Bundoora, VIC 3086, Australia

**Keywords:** Parkinson’s disease, exercise, strength, physical therapy, Powerlifting

## Abstract

**Background:** Although powerlifting has been shown to increase strength and function in some adults, the safety and feasibility of powerlifting exercises modified for people with early onset Parkinson’s disease is not known.

**Methods:** People with early onset Parkinson’s disease (age <50 years) will be invited to engage in eight consecutive weeks of twice-weekly modified powerlifting exercises in a community gymnasium. The primary outcome is the feasibility of powerlifting exercises modified for people with early onset Parkinson’s disease, quantified by measures of safety, adverse events, adherence, access, and cost. Secondary outcomes include motor disability, quality of life, psychosocial status, and strength. Semi-structured interviews with participants and exercise trainers will capture experiences, beliefs, and attitudes towards this type of community-based strength training.

**Discussion:** Powerlifting may be beneficial for those with early onset Parkinson’s disease as it can improve muscle strength, coordination, and balance, and may have neuroprotective effects, potentially slowing down the progression of the disease. However, there is a need to first measure feasibility and safety of modified powerlifting in a small number of individuals before the efficacy can be tested in larger randomised controlled trials.

**Ethics and dissemination:** We will publish the findings in peer-reviewed journals and presentations at conferences. The consumer engagement council will guide communication of findings to individuals with early onset Parkinson’s disease, ensuring information is accessible and actionable for the target audience.

## BACKGROUND

People with Parkinson’s disease who are diagnosed before 50 years of age are described as having early onset Parkinson’s disease^1^. They can face unique challenges such as early onset bradykinesia, dystonia, dyskinesia, and anxiety^2,3^. While Parkinson’s medication is effective for the symptomatic treatment of movement disorders in the early stages following diagnosis, some individuals face troubling motor fluctuations as the disease progresses^4^.

Alongside pharmacological treatment, resistance training is often recommended for people with Parkinson’s disease to improve muscle strength and motor function^5^. Traditional resistance training sessions, which often require high training volumes for benefit, can be time-consuming^6^ and fatiguing^7^. As an alternative, modified powerlifting-style resistance training may be beneficial for people with early onset Parkinson’s disease. Powerlifting is a form of high-intensity strength training which predominantly focusses on three primary exercises: the squat, bench press, and deadlift^8^. It emphasises large multi-joint coordinated movements and can be performed one or two times per week to enhance strength^9,10^. Powerlifting exercises can be tailored to individual abilities by adjusting intensity, range of motion, the use of assistive equipment, and the length and frequency of rest intervals.

The aim of this study is to assess the feasibility and safety of a modified powerlifting-style training intervention at a community gym for people with early onset Parkinson’s disease. Community gyms, along with their trainers, offer a secure environment for supervised strength-focused workouts, which can be challenging to replicate at home.

## METHODS AND ANALYSIS

Ethics approval was obtained from Latrobe University Human Research Ethics Committee (HEC24036) and the trial has been prospectively registered in the Australian New Zealand Clinical Trials Registry (ACTRN1262400044656). People with early onset Parkinson’s disease will be invited to participate in an eight-week, 60-minute prospective, pre-post, feasibility study. The modified powerlifting exercise sessions will be conducted within a community gymnasium to optimise access. The study aligns with the *Strengthening the Reporting of Observational Studies in Epidemiology* (STROBE) checklist and the *Standard Protocol Items: Recommendations for Interventional Trials* (SPIRIT).

### Inclusion and exclusion criteria

Participants must be adults diagnosed with early onset Parkinson’s disease (modified Hoehn & Yahr stage 0-2.5) and able to attend a community gym. They need to be proficient in English and be willing and able to provide informed consent. People will be excluded if they have other neurological disorders, comorbidities impacting gait or impeding safe exercise, orthostatic hypotension, a history of smoking, heart disease, lung conditions, musculoskeletal conditions affecting participation in gym activities, injuries, uncontrolled diabetes, mood disorders, chronic fatigue, cognitive impairment, unmanaged medical or surgical conditions, severe auditory or visual impairment, cancer, obesity, bone disease, receiving deep brain stimulation or using Parkinson’s medication pumps.

Exercise trainers, also referred to as personal trainers, will supervise and oversee the modified powerlifting sessions and offer views on the feasibility of this intervention. To be eligible, exercise trainers must be at least 18 years old, proficient in English, and willing to provide informed consent. They will hold certification as a personal trainer, physiotherapist, or exercise physiologist.

### Recruitment

Participants with early onset Parkinson’s disease will be recruited through flyers distributed in health professional clinics across the Melbourne metropolitan region and via not-for profit agencies. A doctor’s letter will need to be provided advising safety to participate. Exercise trainers from the community gym where the intervention takes place will be recruited through flyers distributed within the gym.

### Informed Consent

After the initial screening process, participants with early onset Parkinson’s disease and the exercise trainers will receive a detailed participant information and consent form. Participants will have ample opportunity to ask questions and address any concerns before signing an informed consent form.

### Consumer Engagement Council

A consumer engagement council (CEC) will gather perspectives, insights, and feedback for the trial. The CEC will be led by a health professional and include people with early onset Parkinson’s disease. The CEC will offer guidance on enhancing recruitment efforts. Following the completion of data collection, the CEC will provide input into interpreting study results from a consumer perspective.

### Exercise Trainer Workshops

The exercise trainers will participate in a “train the trainer” session to credential them in working safely with individuals with early onset Parkinson’s disease. These sessions will cover the characteristics, symptoms, and presentation of early onset Parkinson’s disease and associated safety precautions. This session will also cover how to safely deliver resistance exercises, adapting resistance exercises for people with Parkinson’s disease, and how to safely modify training through personalised prescription, progression, and monitoring.

### Reporting Adverse Events

Exercise trainers will also be instructed to immediately respond to and document any adverse event. The exercise trainers will adhere to their standard operating procedures of their employment workplace for managing adverse events, ensuring prompt first aid and medical assistance if necessary. In the unlikely event of a serious adverse event, such as an injurious fall, the exercise trainers will immediately administer first aid according to standard operating procedures and, if necessary, promptly call an ambulance. The participant’s caregiver or family will also be quickly notified. Serious adverse events will be reported to the La Trobe University Human Ethics Committee within 48 hours.

### Modified Powerlifting Intervention

The powerlifting exercises will be modified to fit the individual abilities of participants. This could include decreasing the exercise intensity and range of motion, performing simpler versions of the main exercises, using assistive equipment where appropriate, or allowing longer or more rest periods to reduce fatigue.

The intervention length is eight weeks, totalling 16 exercise training sessions. Each of the 16 sessions will last for a maximum of one hour and will consist of a structured format. This includes a warm-up phase lasting at least 10 minutes, involving activities such as using a stationary bicycle or rowing machine, but excluding weights. Then the strength training phase will take place for 20-40 minutes. The session will conclude with a cool-down phase on the mat, comprising stretches, yoga, relaxation, or breathing exercises, for at least 10 minutes.

Sessions 1-2 will serve as familiarisation sessions, focusing on orienting participants to the gym environment, safe exercise techniques, gym safety, and education regarding rest periods and rehydration. Exercise trainers will adhere to local policies and procedures, including an exercise screening tool.

Sessions 3-15 will involve the exercise trainers guiding and supervising participants through the warm-up, strength training intervention, and cool-down phases. The exercise trainers will adhere to the evidence-based guidelines provided in the exercise trainer manual developed by our research team. These guidelines were developed based on the resistance exercise recommendations by Martignon et al^11^, ensuring consistency and alignment with established best practices. Exercise trainers will be instructed to modify the exercises to accommodate movement disorders and non-motor symptoms associated with Parkinson’s disease, Parkinson’s medication status, and individual differences. Each participant will have a tailored program adapted for safety.

### Outcome measures

Assessments will take place at the training venue by a registered neurological physiotherapist, during the ON phase of the medication cycle. The primary outcome will be assessing feasibility, measured by recording details around (i) safety, including adverse events, (ii) recruitment/retention rates of people with Parkinson’s disease, including dropouts, (iii) access to physical infrastructure and resources (iv) staffing (v) adherence to session program schedules (vi) cost of delivering the powerlifting programs (vii) participant perceptions on acceptability, benefits and limitations, with interviews conducted with people with Parkinson’s disease and exercise trainers.

Pre- and post-secondary outcome measurements will be conducted during sessions 1 and 16 and will include motor disability (UPDRS, section III), quality of life (EuroQOL 5D VAS), psychosocial functioning (SCOPA-PS) and strength (three-repetition maximum [3RM]). For the 3RM testing, upper body and lower body strength will be assessed using a chest press and leg press machine. The SCOPA-PS will be used to assess how well a person manages both their psychological well-being and their social interactions with others and includes things like mood, coping skills, and relationships.

### Focus Groups

At the completion of the eight-week intervention, participants with Parkinson’s disease will be invited to join a focus group, and exercise trainers will be invited to participate in semi-structured interviews ^12^. These will be facilitated by a member of the research team who will follow interview guides with open-ended questions and prompts. This will be an inductive process using a qualitative descriptive approach to explore the beliefs, experiences, attitudes, and interactions regarding the powerlifting sessions ^13,14^.

### Sample Size

For this pilot feasibility trial, and in-line with previous pilot studies in Parkinson’s disease^15-18^, a convenience sampling method will be used to recruit adults with early onset Parkinson’s disease. A similar number of exercise trainers will also be invited to participate.

### Data Analysis

For the people living with Parkinson’s disease, demographic and clinical characteristics, both continuous and categorical variables, will be reported using descriptive statistics, including numbers (n) and proportions (%) for the participants’ sex, age, height, weight, years living with Parkinson’s disease, and years on Parkinson’s medication(s). Participant height and weight will be used to calculate body mass index (BMI).

We will primarily use descriptive statistics to report on the primary outcome of feasibility, which includes reporting safety, attendance, adherence, and program cost. For the secondary outcomes, paired *t*-tests (two-sided) or Wilcoxon signed-rank tests will be used to assess within-group changes for the continuous variables and the McNemar test will be used for categorical variables. The clinical significance of the results will be based on the magnitude of the Cohen’s d effects size: small (*d*=0.20), medium (*d*=0.50), and large (*d*=0.80)^19^. Quantitative statistical analysis will be conducted using Statistical Package for Social Science (SPSS® v.23 for Windows, IBM), with the significance level set at *p*lJ<□0.05.

Focus group discussions will be summarized trough audio recording using a descriptive approach which will allow for clear information to be obtained when exploring potential improvements of this feasibility study ^20^.

## DISCUSSION

This world-first exercise trial will provide preliminary insight into the safety and feasibility of modified powerlifting for adults with early onset Parkinson’s disease. The findings will guide the design of a future larger randomised controlled trial to test the efficacy of this investigation. Very few exercise studies have reported the feasibility or benefits of community-based strength training for Parkinson’s disease. Most previous strength training trials were on older people with Parkinson’s disease^21^ or people with Parkinson’s disease performing other types of physical activity such as dancing^22^, aquatic therapy^23^ or boxing^24^. Given the potential for high-intensity exercise to modify the rate of disease progression in Parkinson’s disease^25, 26^ the results have the potential to positively impact health and wellbeing.

## Data Availability

All data produced from this feasibility trial will be made available upon reasonable request to the authors.

## Contributors

All authors designed the study, formulated the research questions, and contributed to this manuscript.

## Patient consent for publication

Not required.

## References

1. Bloem BR, Okun MS and Klein C. Parkinson’s disease. Lancet 2021; 397: 2284–2303. 20210410. DOI: 10.1016/s0140-6736(21)00218-x.

2. Mehanna R and Jankovic J. Young-onset Parkinson’s disease: Its unique features and their impact on quality of life. Parkinsonism & Related Disorders 2019; 65: 39–48.

3. Mehanna R, Moore S, Hou JG, et al. Comparing clinical features of young onset, middle onset and late onset Parkinson’s disease. Parkinsonism & related disorders 2014; 20: 530–534.

4. Freitas ME, Hess CW and Fox SH. Motor Complications of Dopaminergic Medications in Parkinson’s Disease. Semin Neurol 2017; 37: 147-157. 20170516. DOI: 10.1055/s-0037-1602423.

5. Yang X and Wang Z. Effectiveness of Progressive Resistance Training in Parkinson’s Disease: A Systematic Review and Meta-Analysis. Eur Neurol 2023; 86: 25-33. Article. DOI: 10.1159/000527029.

6. Iversen VM, Norum M, Schoenfeld BJ, et al. No time to lift? Designing time-efficient training programs for strength and hypertrophy: a narrative review. Sports Medicine 2021; 51: 2079–2095.

7. Barahona-Fuentes GD, Ojeda ÁH and Jerez-Mayorga D. Effects of different methods of strength training on indicators of muscle fatigue during and after strength training: A systematic review. Motriz: Revista de Educação Física 2020; 26.

8. Latella C, Teo W-P, Spathis J, et al. Long-term strength adaptation: A 15-year analysis of powerlifting athletes. J Strength Cond Res 2020; 34: 2412.

9. Fyfe JJ, Hamilton DL and Daly RM. Minimal-Dose Resistance Training for Improving Muscle Mass, Strength, and Function: A Narrative Review of Current Evidence and Practical Considerations. Sports Med 2022; 52: 463-479. 2021/11/26. DOI: 10.1007/s40279-021-01605-8.

10. Behm DG, Granacher U, Warneke K, et al. Minimalist Training: Is Lower Dosage or Intensity Resistance Training Effective to Improve Physical Fitness? A Narrative Review. Sports Medicine 2024; 54: 289–302. DOI: 10.1007/s40279-023-01949-3.

11. Martignon C, Pedrinolla A, Ruzzante F, et al. Guidelines on exercise testing and prescription for patients at different stages of Parkinson’s disease. Aging Clinical and Experimental Research 2021; 33: 221–246. DOI: 10.1007/s40520-020-01612-1.

12. Braun V and Clarke V. Using thematic analysis in psychology. Qualitative Research in Psychology 2006; 3: 77–101. DOI: 10.1191/1478088706qp063oa.

13. Bradshaw C, Atkinson S and Doody O. Employing a Qualitative Description Approach in Health Care Research. Global Qualitative Nursing Research 2017; 4: 2333393617742282. DOI: 10.1177/2333393617742282.

14. Doyle L, McCabe C, Keogh B, et al. An overview of the qualitative descriptive design within nursing research. J Res Nurs 2020; 25: 443-455. 20191218. DOI: 10.1177/1744987119880234.

15. Kwok JYY, Lee JJ, Choi EPH, et al. Stay mindfully active during the coronavirus pandemic: a feasibility study of mHealth-delivered mindfulness yoga program for people with Parkinson’s disease. BMC Complementary Medicine and Therapies 2022; 22: 37.

16. Lihala S, Mitra S, Neogy S, et al. Dance movement therapy in rehabilitation of Parkinson’s disease–A feasibility study. Journal of bodywork and movement therapies 2021; 26: 12–17.

17. Morris ME, Slade SC, Wittwer JE, et al. Online dance therapy for people with Parkinson’s disease: feasibility and impact on consumer engagement. Neurorehabilitation and Neural Repair 2021; 35: 1076–1087.

18. Shanahan J, Morris ME, Bhriain ON, et al. Is Irish set dancing feasible for people with Parkinson’s disease in Ireland? Complementary therapies in clinical practice 2015; 21: 47–51.

19. Cohen J. Statistical power analysis for the behavioral sciences. Routledge, 2013.

20. Sullivan-Bolyai S, Bova C and Harper D. Developing and refining interventions in persons with health disparities: the use of qualitative description. Nurs Outlook 2005; 53: 127–133. DOI: 10.1016/j.outlook.2005.03.005.

21. Gamborg M, Hvid LG, Dalgas U, et al. Parkinson’s disease and intensive exercise therapy—An updated systematic review and meta□analysis. Acta Neurologica Scandinavica 2022; 145: 504–528.

22. Shanahan J, Morris ME, Bhriain ON, et al. Dancing for Parkinson disease: a randomized trial of Irish set dancing compared with usual care. Archives of physical medicine and rehabilitation 2017; 98: 1744–1751.

23. Carroll LM, Volpe D, Morris ME, et al. Aquatic exercise therapy for people with Parkinson disease: a randomized controlled trial. Archives of physical medicine and rehabilitation 2017; 98: 631–638.

24. Combs SA, Diehl MD, Chrzastowski C, et al. Community-based group exercise for persons with Parkinson disease: a randomized controlled trial. NeuroRehabilitation 2013; 32: 117–124.

25. Helgerud J, Thomsen SN, Hoff J, et al. Maximal strength training in patients with Parkinson’s disease: impact on efferent neural drive, force-generating capacity, and functional performance. Journal of Applied Physiology 2020; 129: 683–690.

26. Dibble LE, Hale TF, Marcus RL, et al. High intensity eccentric resistance training decreases bradykinesia and improves quality of life in persons with Parkinson’s disease: a preliminary study. Parkinsonism & related disorders 2009; 15: 752–757.

